# Bayesian Inference of Nosocomial *Methicillin-resistant Staphylococcus aureus* Transmission Rates in an Urban Safety-Net Hospital

**DOI:** 10.1101/2024.12.18.24319252

**Authors:** Kiel Corkran, Majid Bani-Yaghoub, Gary Sutkin, Arash Arjmand, Susanna Paschal

## Abstract

Methicillin-resistant *Staphylococcus aureus* (MRSA) is a strain of *Staphylococcus aureus* that poses significant challenges in treatment and infection control within healthcare settings. Recent research suggests that the incidence of healthcare-associated MRSA (HA-MRSA) is higher among patients treated in safety-net hospitals compared to those in non-safety-net hospitals.

This study aimed to identify HA-MRSA transmission patterns across various nursing units of a safety-net hospital to improve to enhance patient outcomes and facilitate the implementation of targeted infection control measures.

A retrospective analysis was conducted using surveillance data from 2019 to 2023. A compartmental disease model was applied to estimate MRSA transmission rates and basic reproduction number (*R*_0_) for each nursing unit of an urban, multicenter safety-net hospital before and during the COVID-19 pandemic. Posterior probability distributions for transmission, isolation, and hospital discharge rates were computed using the Delayed Rejection Adaptive Metropolis (DRAM) Bayesian algorithm.

Analysis of 187,040 patient records revealed that inpatient nursing units exhibited the highest MRSA transmission rates in three out of the five years studied. Notable transmission rates were observed in certain inpatient and progressive care units (0.55 per individual per month; 0.018 per individual per day) and the surgical ICU (0.44 per individual per month; 0.015 per individual per day). In contrast, the Nursery NICU and Medical ICU had the lowest transmission rates. Although MRSA transmission rates significantly declined across all units in 2021, these rates rebounded to pre-pandemic levels in subsequent years. Notably, outbreaks emerged in units such as ICUs and progressive care units that had not experienced prior MRSA outbreaks since 2019.

While MRSA transmission significantly declined during the initial phase of the pandemic, the pathogen reestablished itself in later years. These findings highlight the need for sustained resources and adaptive infection control strategies to reduce the incidence of HA-MRSA in safety-net hospitals.

## Introduction

*Methicillin-resistant Staphylococcus aureus (MRSA)* is a type of *Staphylococcus aureus* bacteria resistant to multiple antibiotics. While S. aureus is commonly found on the skin or in the nose of approximately 30% of the population [1], MRSA infections can cause severe illness or even death if untreated [2]. High-risk groups include athletes, students, military personnel, individuals receiving inpatient care, and those with surgeries or medical devices [2]. MRSA symptoms vary based on the infection site and source, whether from healthcare or community settings. Common symptoms include swollen, painful skin infections, pneumonia, urinary tract infections, bloodstream infections, and infected surgical wounds [1, 3, 4]. In hospitals, contaminated surfaces and medical devices, often originating from colonized patients or staff, are primary sources [1]. In communities, factors include group living, recent illegal drug use, poor hygiene, and contact sports [5]. Animals can also serve as reservoirs for MRSA [6]. Treatment options range from chemotherapeutics and natural drugs to multi-drug strategies, bacteriophage-antibiotic combinations, and ongoing efforts to develop anti-MRSA vaccines [7].

Globally, MRSA accounts for 13–74% of S. aureus infections, with prevalence and incidence rates varying. Recent studies report hospital incidence at 22.58% and non-hospital incidence at 11.59% [8, 9]. The CDC estimates MRSA causes over 70,000 severe infections and 9,000 deaths annually in the U.S. [11, 13]. MRSA infections can be classified into healthcare-associated (HA-MRSA), community-associated (CA-MRSA), and livestock-associated (LA-MRSA) categories based on their source. HA-MRSA is associated with healthcare exposure within a year before culture, such as surgery, hospitalization, hemodialysis, or residence in a long-term care facility, or with hospitalization lasting more than 4 days at the time of culture [12]. In contrast, CA-MRSA affects individuals with no history of recent healthcare exposure, invasive medical devices, dialysis, or prior MRSA infection or colonization within the year preceding the culture [12].

Research on MRSA transmission has examined intervention strategies such as hand hygiene, screening, and workload adjustments. Increasing handwashing rates during contact with bodily fluids and screening patients at admission were among the most effective strategies for reducing MRSA transmission [31, 32]. Economic analyses have highlighted the costs associated with MRSA infections and the cost-effectiveness of various screening methods [33]. Scandinavian and Dutch “search-and-destroy” strategies effectively lowered MRSA remission rates [34]. Patient readmissions were shown to significantly contribute to HA-MRSA, with a 44.2% likelihood of readmitted patients being infected [35]. In military medical facilities, baseline MRSA acquisition rates were analyzed, with MRSA patients averaging 17.7 hospital days compared to 5.3 for uninfected patients [36]. A Bayesian model of over 230 VA hospitals and nursing homes found that MRSA transmission rates were four times higher in hospitals than in nursing homes [14].

A significant healthcare debate concerns whether reducing HA-MRSA rates effectively lowers the overall healthcare-associated infection (HAI) burden or merely results in its replacement by other pathogens, leaving the total disease burden unchanged. For instance, a recent study [44] analyzed data from over 1 million patients across 51 acute care facilities in the U.S. Using a threshold model, the study demonstrated that MRSA contributes additively to the total HAI burden. The findings suggest that reducing HA-MRSA rates can decrease the overall nosocomial infection rate, highlighting the indicator role of MRSA and its distinct public health niche.

Despite numerous studies on modeling HA-MRSA transmission dynamics [30-36], limited research has addressed HA-MRSA transmission in safety-net hospitals across different nursing units. This focus is critical, as recent evidence indicates disproportionately higher rates of HA-MRSA in safety-net hospitals compared to non-safety-net hospitals, partly due to socioeconomic disparities and inequities in healthcare resources [46, 47, 49]. Moreover, MRSA among minoritized racial, ethnic, and language groups, even after adjusting for other known risk factors [45, 48, 49]. To address this gap, the present study analyzes 2019–2023 surveillance data from a safety-net hospital to model the transmission dynamics of HA-MRSA using Bayesian inference techniques. By identifying significant transmission patterns across various nursing units, this research aims to inform targeted interventions and equitable healthcare strategies for reducing HA-MRSA in various nursing units. Ultimately, this research seeks to promote more equitable healthcare approaches that reduce HA-MRSA transmission, improve patient outcomes, and address the underlying systemic factors that contribute to the disproportionate burden of MRSA in safety-net settings.

## Materials and Methods

### Study Setting

This study was conducted at University Health in Kansas City, Missouri, the primary academic medical center for the University of Missouri-Kansas City (UMKC) School of Medicine. As the nonprofit entity succeeding Kansas City’s public hospitals, University Health fulfills a critical role as an urban safety-net provider. Approximately half of its patients are from historically marginalized populations, with the majority lacking adequate insurance coverage. Medicaid and Medicare collectively represent 72% of patient visits, 59.7% of discharges, and 63.3% of net patient revenue, while commercial insurance accounts for a comparatively small share. In 2021, University Health delivered nearly $140 million in uncompensated care, highlighting its fundamental role in mitigating healthcare disparities across the region [46, 51]. The organization operates two principal facilities: Truman Medical Center (TMC), housing 258 staffed beds, and Lakewood Medical Center (LMC), with 110 staffed beds [17, 18].

### Study Population

The dataset provided by University Health Kansas City includes comprehensive records of patient admissions, discharges, and infections for individuals hospitalized between 2019 and 2023. Over this period, 789 patients were hospitalized for at least four days and tested positive for MRSA, accounting for 39.8% of all healthcare-associated infections (HAIs) recorded during the study period (see Table S1 and S2 in the supplementary document).

To evaluate MRSA infection trends, cases were clustered across 13 nursing units categorized into inpatient and non-inpatient units to identify variations in infection patterns and capabilities. The inpatient units included 3^rd^ Floor Blue Unit (IU1), Gold Unit (IU2), and Red Unit (IU3), along with 4^th^ Floor Blue Unit (IU4), Green Unit (IU5), Gold Unit (IU6), and Red Unit (IU7). Additionally, 5^th^ Floor North Unit (IU8) was part of this group. The non-inpatient units comprised critical care units such as the Cardiac Care Unit (CCU), Neonatal Intensive Care Unit (NICU), Progressive Care Unit (PCU), Intensive Care Unit (ICU), and Mental Health (MH). This classification provided a detailed framework for analyzing MRSA infection trends across different types of nursing units. See Table 1 for summary statistics.

**Table 1.** Summary monthly statistics of MRSA incidents in different locations of the Lakewood and Truman Medical Center Nursing Units during 2019-2023.

|  | CCU | IU1 | IU2 | IU3 | IU4 | IU5 | IU6 | IU7 | IU8 | MS | NICU | PCU | ICU | MH |
| --- | --- | --- | --- | --- | --- | --- | --- | --- | --- | --- | --- | --- | --- | --- |
| Mean | 0.25 | 1.2 | 2.7 | 1.2 | 2 | 1.1 | 0.8 | 1.6 | 0.1 | 0.8 | 0.06 | 0.38 | 0.26 | 0 |
| Median | 0 | 0 | 2 | 1 | 2 | 1 | 0 | 2 | 0 | 0 | 0 | 0 | 0 | 0 |
| Max | 2 | 6 | 9 | 3 | 7 | 3 | 4 | 4 | 1 | 5 | 2 | 3 | 3 | 1 |
| Sum | 15 | 64 | 163 | 69 | 118 | 66 | 48 | 97 | 3 | 48 | 4 | 23 | 16 | 2 |

There are 8 inpatient units denoted by IU1-8, Cardiac Care Unit (CCU), Neonatal Intensive Care Unit (NICU), Progressive Care Unit (PCU), Intensive Care Unit (ICU), Mental Health (MH).

### The UCQR Model

Figure 1 shows a schematic representation of the proposed Uncolonized-Colonized-Quarantined-Recovered (UCQR) model. Uncolonized patients admitted to the hospital may be exposed to MRSA from colonized individuals, including healthcare workers or other patients, and subsequently become colonized with MRSA. Colonized patients who test positive for MRSA or develop MRSA symptoms will be isolated in a private room to prevent transmission of the infection. Asymptomatic colonized patients who do not require isolation may be discharged after completing their treatment, although infection control measures will still be implemented to mitigate the risk of MRSA transmission.

**Figure 1:**
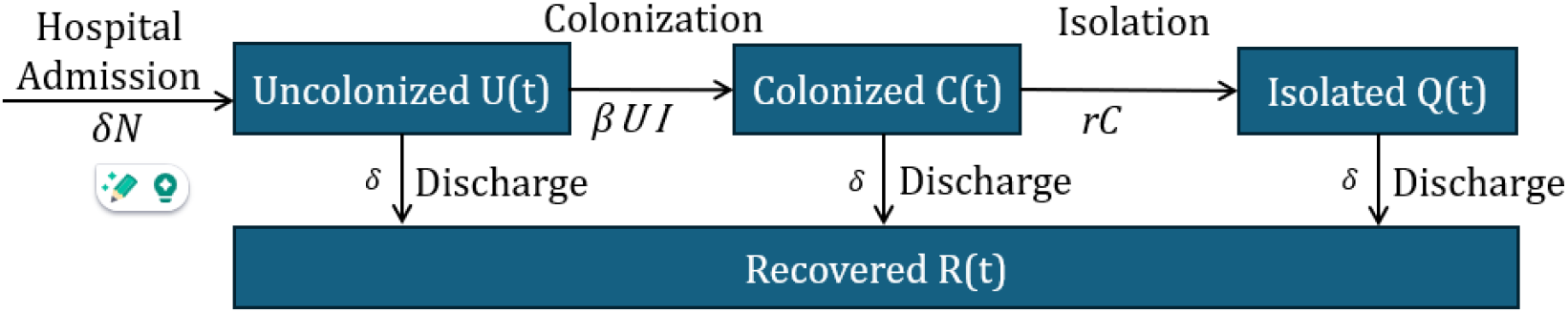
Schematic representation of the Uncolonized-Colonized-Quarantined-Recovered (UCQR) Model. From any compartment, a patient can be discharged, including those MRSA colonized but unidentified.

The following assumptions were made in the model construction: (1) most colonized patients are identified and quarantined, including those upon admission; (2) MRSA colonization precedes the onset of symptoms; (3) MRSA-colonized patients primarily transmit the infection indirectly to other patients, with healthcare workers and contaminated fomites serving as common intermediaries; (4) Patients who are isolated due to MRSA have a negligible risk of directly transmitting MRSA to other patients.

Let *U*(*t*), *C*(*t*), *Q*(*t*), and *R*(*t*) be the number of uncolonized, colonized, quarantined, and recovered individuals at time *t*, respectively. The UCQR model is then formulated as a system of Ordinary Differential Equations (ODEs) as follows:

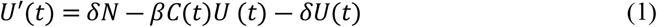

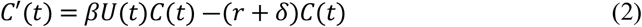

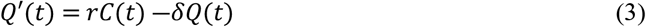

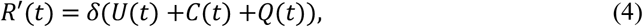

where *β* is the average transmission rate, *r* is the isolation rate, *δN* is the hospital admission rate at time *t*, and *δ* the hospital discharge rate.

Using the records provided by University Health, we established baseline values *β*_0_, *r*_0_, *δ*_0_ for the unknown parameters. Baseline values are in section 2 of the supplementary (see Table S3).

### Bayesian Algorithm for Parameter Estimation

We used the Bayesian approach “*Delayed-Rejection Adaptive-Metropolis (DRAM) algorithm*” to find the posterior density that best reflects the distribution of the unknown parameters *q* = [*β, δ*,*r*] of the UCQR Model. DRAM is a widely used and accepted Bayesian method for statistical inference that combines Delayed Rejection and Adaptive Metropolis algorithms [10, 19, 21, 22]. Below is an outline of these methods.

Let (Ω,*F*,*P*) be a probability space, where Ω is the set of elementary events (sample space), *F* a *σ*-algebra of events, and P a probability measure. Let *π*_0_(*q*), *π*(*q│y*), and *π*(*y│q*) be the probability density functions of the prior, posterior, and sampling distributions, respectively. Using Bayes’ theorem, we have

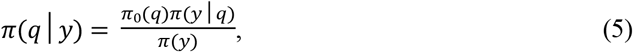

where

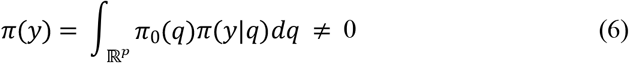

The density *π*(*y│q*), commonly referred to as the likelihood function, incorporates information from measurement data to update prior knowledge. The objective of Bayesian inversion is to determine the posterior probability density function *π*(*q│y*), which quantifies the uncertainty associated with the model parameter set *q* = [*β, δ*,*r*], informed by the observed data *y*, which is the monthly incidence of MRSA in different nursing units. Nevertheless, calculating the integral in equation (6) is computationally costly [10, 21].

The Metropolis-Hastings (MH) algorithm [10, 21, 22], a Markov Chain Monte Carlo (MCMC) method, facilitates Bayesian parameter estimation without requiring the computation of integral (6). MH constructs a Markov chain by accepting or rejecting proposed parameter values based on the posterior distribution. In particular, the new state *q*_*k*_ is constructed based on the previous state *q*_*k*–1_ and a new proposed value *q*^∗^ obtained by the proposal density function 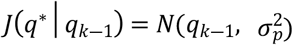 where *σ*_*p*_ is the proposal covariance. Then the following acceptance ratio is used to accept *q*^∗^ or reject it and keep the old value *q*_*k*–1_.

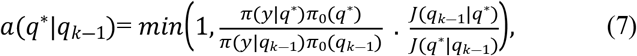

where the likelihood distribution is defined by

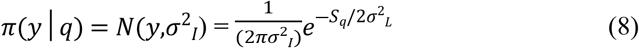

with the likelihood covariance, *σ*_*p*_ the residual sum squares 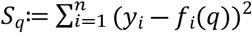 and *f*(*q*) is the parameter-dependent UCQR model response. Adaptive algorithms, such as the Delayed Rejection Adaptive Metropolis (DRAM), address the inefficiencies of the trial-and-error proposal searches by optimizing proposal states and refining the rejection process [10, 24, 25, 26]. If an initial proposal *q*^∗^ is rejected, the delayed rejection mechanism generates an alternative candidate *q*^∗∗^ using density function

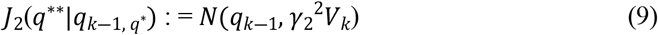

where

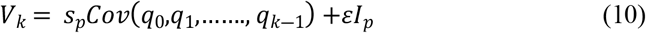

*ε* and *S*_*p*_ are constants and *I*_*p*_ *is* the *p*-dimensional identity matrix to ensure that *V*_*k*_ is invertible. The Adaptive Metropolis (AM) algorithm employs a global adaptive strategy based on a recursive relation

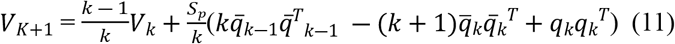

with the sample mean

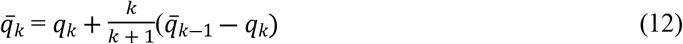

to update the proposal covariance matrix using the Gaussian proposal centered at *q*_*k*_ and update the chain covariance matrix at the *k*-th step.

## Results

### MRSA Transmission Rates

We obtained the posterior distributions of the parameters *q* = [*β, δ*,*r*] using the DRAM algorithm for Bayesian inversion. Sample plots of the estimated distribution of model parameters *β, δ*,*r* can be found in Section 3 of the supplementary Document (See Figures S1-S8). Table 2 presents the estimated confidence intervals and means of the Markov chains for MRSA transmission rates *β* across nursing units from 2019 to 2023. Note that all rates *β* values per individual per month. As described below, the space-time pattern of MRSA transmission rates *β* reveals significant variability across nursing units and years (2019–2023).

**Table 2:** Space-time patterns of mean MRSA transmission rates and ( 95% confidence intervals) in different nursing units of the University Health during 2019-2023. The gray boxes represent the spikes. The estimated 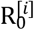 values are shown inside brackets.

| $\beta$ | Nursing Unit | Year 2019 | Year 2020 | Year 2021 | Year 2022 | Year 2023 |
| --- | --- | --- | --- | --- | --- | --- |
| $\beta_1$ | Cardiac Care Center ($N = 8$) | .0030<br>(.0030, .0031) | 0 | 0 | .0030<br>(.0029, .0030) | .0033<br>(.0032, .0033) |
| $\beta_2$ | Blue Inpatient FL3 ($N = 7$) | .0032<br>(.0031, .0032) | .0038<br>(.0037, .0039) | .0035<br>(.0034, .0035) | 0 | .0033<br>(.0032, .0033) |
| $\beta_3$ | Gold Inpatient FL3 ($N = 10$) | .5489 [4.89]<br>(.5437, .5541) | .0032<br>(.0031, .0032) | .0033<br>(.0033, .0034) | .0032<br>(.0032, .0033) | .0032<br>(.0032, .0033) |
| $\beta_4$ | Inpatient Red FL3 ($N = 7$) | .0034<br>(.0034, .0035) | .0030<br>(.0029, .0030) | .0034<br>(.0033, .0035) | .0033<br>(.0032, .0034) | .0031<br>(.0030, .0031) |
| $\beta_5$ | Inpatient Blue FL4 ($N = 7$) | .4370 [2.95]<br>(.4308, .4426) | .4347 [2.91]<br>(.4291, .4403) | .0030<br>(.0030, .0031) | .0030<br>(.0029, .0030) | .0031<br>(.00031, .0032) |
| $\beta_6$ | Inpatient Green FL4 ($N = 12$) | .0031<br>(.0030, .0032) | .0032<br>(.0031, .0032) | 0.0032<br>(.0031, .0032) | .0030<br>(.0030, .0031) | .0034<br>(.0034, .0035) |
| $\beta_7$ | Inpatient Gold FL4 ($N = 10$) | 0 | 0 | 0 | 0 | .4241 [4.10]<br>(.4180, .4298) |
| $\beta_8$ | Inpatient Red FL4 ($N = 7$) | .0030<br>(.0029, .0030) | 0 | .0031<br>(.0031, .0032) | .4536 [3.09]<br>(.4476, .4597) | .0031<br>(.0030, .0031) |
| $\beta_9$ | Nursery NICU ($N = 18$) | 0.0031<br>(.0031, .0032) | 0.0032<br>(.0031, .0033) | 0 | 0 | 0 |
| $\beta_{10}$ | Progressive Care Unit ($N = 12$) | .5491 [5.62]<br>(.5435, .5546) | .0029<br>(.0029, .0030) | .0031<br>(.0030, .0032) | .4166 [4.77]<br>(.4110, .4223) | .0031<br>(.0030, .0032) |
| $\beta_{11}$ | Surgery ICU ($N = 16$) | .0029<br>(.0029, .0030) | .0031<br>(.0030, .0031) | .0031<br>(.0030, .0032) | .4444 [6.51]<br>(.4388, .4501) | .0031<br>(.0030, .0031) |
| $\beta_{12}$ | Inpatient North FL5 ($N = 6$) | 0 | 0 | .0031<br>(.0031, .0032) | .4697 [2.72]<br>(.4637, .4755) | .0031<br>(.0030, .0031) |
| $\beta_{13}$ | Medical ICU II ($N = 50$) | 0 | 0 | .0034<br>(.0033, .0035) | 0 | 0 |

*All rates $\beta$ values per individual per month*

Some units, such as the Cardiac Care Center, 3^rd^ Floor Inpatient Blue Unit, and the Nursery NICU, consistently exhibit low transmission rates (*β* ≈ 0.003) with only minor fluctuations over the years. In contrast, certain units demonstrate persistently high transmission rates, notably the Progressive Care Unit, which experienced elevated rates in 2019 (*β* = 0.5491) and again in 2022 (*β* = 0.4166). Similarly, Floor 4 Inpatient Blue Unit maintained high rates in 2019 and 2020 (*β* ≈ 0.4370) before declining sharply in later years.

Other nursing units show emerging or isolated spikes in transmission. For example, Floor 4 Inpatient Red Unit experienced a dramatic peak in 2022 (*β* = 0.4536), while Floor 4 Inpatient Gold Unit exhibited a sudden spike in 2023 (*β* = 0.4241) after years of insignificant MRSA transmission. A similar trend is observed in Floor 5 Inpatient North Unit, which substantially increased in 2022 (*β* = 0.4697) following zero rates in prior years. Conversely, some units, such as the Nursery NICU and Medical ICU II, show minimal activity, with sporadic low transmission rates (*β* ≈ 0.003).

Over time, the temporal pattern indicates both stability and fluctuation depending on the unit. Some units, like the Cardiac Care Center and Floor 3 Red Section, maintain low and consistent transmission rates throughout the study period. Others, such as Floor 3 Blue Unit and Floor 4 Inpatient Green Unit, exhibit moderate fluctuations, alternating between slightly elevated (*β* ≈ 0.003) and zero transmission. Certain units display episodic peaks, with high transmission rates appearing sporadically. For instance, in 2019, elevated activity was observed in the Progressive Care Unit and Floor 4 Blue Unit, while in 2022, spikes were recorded in Floor 4 Red Unit and Floor 5 North Unit. The Floor 4 Gold Unit stands out with its significant peak in 2023 after years of inactivity. These findings highlight the heterogeneity of MRSA transmission within the hospital and can facilitate the implementation of targeted infection control measures. Specific units, such as the Progressive Care Unit, appear particularly prone to repeated outbreaks, while others experience episodic spikes potentially linked to localized factors such as patient population changes or procedural shifts. The consistently low rates in certain units suggest effective infection control measures are in place. Tailored interventions targeting high-risk units and addressing periods of increased transmission are crucial for managing MRSA within this setting.

### Basic Reproduction Number

The basic reproduction number *R*_0_ represents the expected number of secondary cases generated by an infected individual in a fully uncolonized population [37]. If *R*_0_ exceeds one in a specific nursing unit, MRSA transmission may become endemic within that unit. Conversely, if *R*_0_ is below one, MRSA infections will decline, leading the unit to an MRSA-free equilibrium. We estimated *R*_0_ using the next-generation matrix approach [37], with inputs derived directly from the estimated *q* = [*β, δ*,*r*] of the UCQR model, following the formula *R*_0_= *Nβ/*( *δ* + *r*). Although more advanced methods such as those described in [15] are available for estimating basic reproduction number 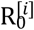 for each nursing unit *i*, their implementation requires additional data that is not currently available. Here, we used the formula 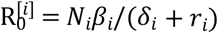, assuming that the number of patients *N*_*i*_ in unit *i* is mainly constant. The estimated values of 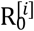 have been indicated in Table 2 for nursing units with high MRSA transmission rates, which vary between 2.72 and 6.51. For all other units, the estimated 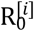 values were less than 1.

### Isolation Rates

Using the outputs of DARM simulations, we calculated the mean isolation rates *r* per month for each nursing unit, complementing the transmission rate analysis. These mean values, along with their 95% confidence intervals, are detailed in Supplementary Table S4. The highest mean isolation rates were observed in inpatient nursing units on floors three and four at both hospitals, except in 2019 and 2021, when non-inpatient nursing units showed higher isolation rates. For instance, in 2021, the surgery ICU and medical ICU II had the highest isolation rates of 0.5926 and 0.5700, respectively, with other units slightly lower.

### Hospital Discharge Rates

Mean monthly hospital discharge rates *δ* were calculated for each nursing unit using the DRAM algorithm as part of the Bayesian inversion solution set. As with previous analyses, 95% confidence intervals were computed to validate the discharge rate estimates. These mean discharge rates and their confidence intervals are presented in Supplementary Table S5. Unlike transmission and isolation rates, the highest discharge rates were observed in non-inpatient units, including the Nursery NICU, Progressive Care Unit, Surgery ICU, and Medical ICU II. Discharge rates generally followed a time-location pattern, showing a slight annual increase until a threshold was reached, after which rates began to decline. This cyclical pattern occurred multiple times in certain units, such as the inpatient blue unit on floor 3, where discharge rates increased from 0.5574 in 2020 before fluctuating in subsequent years. Overall, mean discharge rates ranged from 0.4800 to 0.5700 between 2019 and 2023.

## Discussion

Very few studies have focused on modeling the transmission dynamics of HA-MRSA in safety net hospitals across different nursing units. To fill this gap, the present study utilized 2019-2023 surveillance data of a safety-net hospital to model transmission dynamics of HA-MRSA before and during the COVID-19 pandemic. The Centers for Disease Control and Prevention (CDC) reports that bacterial antimicrobial-resistant hospital-onset infections increased by 20% during the COVID-19 pandemic compared to pre-pandemic levels, peaking in 2021 [40]. By 2022, rates for most pathogens, except MRSA, remained elevated. MRSA rates fluctuated during 2019–2022 [40], with some studies noting increased transmission during the pandemic [42, 43]. As shown in Table 2, MRSA transmission significantly declined during the initial phase of the pandemic, but the pathogen reestablished itself in nursing units such as ICUs and progressive care units.

MRSA transmission rates observed in this study were 0.55 per individual per month (0.018 per individual per day) in inpatient and progressive care units and 0.44 per individual per month (0.015 per individual per day) in the surgical ICU. These rates align with findings from other hospitals and nursing homes [14, 33, 46, 49]. A study conducted in high-admission hospitals in the Netherlands reported an average transmission rate of 0.30, suggesting that MRSA spread was largely under control during the study period [39]. In comparison, the mean transmission rates from Table 2 of this study indicate moderate infection control, though occasional spikes highlight areas for improvement. Another study, employing a methodology similar to the DRAM, reported transmission rates ranging from 0.89 to 0.56, influenced by variations in screening test sensitivity and result reporting times [36]. It should be noted that University Health in Kansas City provides care to underserved populations, making it a valuable setting for further research. Future studies could investigate whether MRSA transmission disproportionately affects specific ethnic and social groups, as was observed with the unequal impact of COVID-19 on certain communities [23, 38].

Several factors may explain the higher MRSA transmission rates observed in inpatient units (Table 2). MRSA spreads through direct contact with open wounds, contaminated surfaces, or via patient-healthcare worker interactions. Inpatient units, where patient-healthcare worker contact rates are higher, inherently present greater opportunities for transmission. Additionally, inpatient floors often accommodate both short-term and long-term care, resulting in higher patient turnover than non-inpatient units, which typically involve shorter hospital stays [32]. Variations in patient flow [16] over time within these 13 units may also account for the transmission spikes observed in the time-location patterns. Similarly, factors affecting patient flow could also contribute to HA-MRSA transmission dynamics [16]. Following the declaration of the COVID-19 pandemic in March 2020, hospital admissions dropped sharply, with some facilities operating at less than 50% capacity [52]. This decrease was partly due to suspending elective surgeries and non-critical services [52]. Reduced admission volumes likely led to fewer MRSA-colonized patients and lower opportunities for transmission. Consequently, the limited influx of colonized and non-colonized patients may have resulted in lower MRSA transmission rates during this period.

It is important to note that the proposed model (1)-(4) carries a number of limitations. A key limitation is the exclusion of explicit compartments for healthcare workers and contaminated fomite density. This simplification, necessitated by insufficient data from University Health, overlooks MRSA infections caused by agents such as contaminated equipment or healthcare workers moving between nursing units. Another limitation is that the model lacks consideration of comorbidities, which could influence the transmission and severity of HA-MRSA infections. Nonetheless, recent studies [44] suggest that HA-MRSA is a biomarker for overall trends in healthcare-associated infections (HAIs). Hence, a decline in MRSA incidence does not result in the emergence of alternative pathogens but rather leads to a substantial reduction in the overall burden of HAIs. The third limitation of the model is its failure to account for community-associated MRSA infections brought into the hospital by patients with short stays (less than 4 days). The pathogen load and interactions between these patients and longer-stay patients could influence the overall MRSA transmission dynamics within the safety-net hospital. Additionally, the Bayesian methodology employed here relies on specific prior distributions for the unknown parameter set (*q*) to ensure proper inversion. Inadequate choice of prior distributions can increase the risk of errors in posterior calculations. Further enhancements to the Bayesian framework will include more adaptive mechanisms in DRAM, enabling it to handle complex prior distributions, such as non-parametric, tail-heavy, or mixture distributions, with improved robustness.

Despite these limitations, the present study is the first step toward a comprehensive analysis of HA-MRSA transmission dynamics in safety-net hospitals. Future research should focus on refining such models by incorporating additional variables to enhance their accuracy and predictive capabilities. Additionally, exploring the impact of different infection control strategies tailored to safety-net hospitals can be essential in mitigating the spread of HA-MRSA and improving patient outcomes.

## Data Availability

To replicate this study with a hypothetical dataset, researchers can download and run the source codes at the following GitHub link:,https://github.com/Corkran1/UHKC-MRSA-Transmission-Model-FIies.

## Model Implementation and Reproducibility

To replicate this study with a hypothetical dataset, researchers can download and run the source codes at the following GitHub link:,https://github.com/Corkran1/UHKC-MRSA-Transmission-Model-FIies. Detailed instructions on how to run the codes and how to interpret the outputs can be found in the supplementary Word document entitled “Instructions to Run Simulations”.

## Ethical approval

The Institutional Review Board (IRB) of the University of Missouri Kansas City approved our study protocol in 2023 (IRB Project Number 2094337). All methods were carried out in accordance with relevant guidelines and regulations.

## Author Contribution

KC contributed to coding and simulations, formal analysis, validation, and manuscript preparation. MB was responsible for conceptualization, developing methodology, data curation, formal analysis, supervision, review and editing, as well as funding acquisition. GS handled project administration, funding acquisition, and review and editing. AA participated in the review and editing of the manuscript, while SP provided resources and contributed to the review and editing process.

## Acknowledgment

This study was supported by the Center for Disease Control and Prevention under grant number 5U01CK000671-03.

